# Real world data on Solitary Plasmacytoma from eastern India- highlighting favorable trends in outcome

**DOI:** 10.64898/2026.04.15.26350956

**Authors:** Dibakar Podder, Himadri Sonowal, Saswata Saha, Bikash Shah, Shouriyo Ghosh, Jeevan Kumar, Arijit Nag, Debranjani Chattyopadhyay, Rizwan Javed, Ashish Rath, Subhosmito Chakraborty, Mayur Parihar, Lateef Zameer, Rimpa Basu Achari, Reena Nair

## Abstract

**Introduction:** Solitary plasmacytomas (SP) are rare neoplasm of localised proliferation of clonal plasma cells. It can be classified based on site of involvement and bone marrow involvement. It is an indolent disease in the majority of patients. Primary modality of treatment is radiotherapy and surgical excision.

**Materials and methods:** This was a retrospective audit of SP who were treated and followed up at a tertiary care center in eastern India from January 2012 to December 2025. Patients who has solitary plasma cytoma with more than 10% plasma cells, POEMS syndrome, have been excluded from analysis.

**Results:** We identified 46 patients of SP. The median age of the studied population was 53 years (23-75 years). Males were more commonly affected than females (M:F=2.2:1). Most common chief complaints were bony pain (67.4%). SBP was seen in 39 (84.8%) cases whereas SEP was seen in 7 (15.2%) cases. Vertebra was the most common site of involvement (61.4%). Median M band concentration 0.24 g/dL (0.1 to 1.95 gm/dL). IgG was the most common isotype accounting for 60.6% cases. Six cases (13%) had minimal bone marrow involvement. The majority of the patients received local radiotherapy (89.1%). With a median follow up of 5.4 years (95% CI: 1.8 – 9.0), median OS was not reached, median PFS was 9.22 years (95% CI: 5.8-12.6), median time to next treatment (TTNT) was 9.86 years (95% CI: 6.8 – 12.9).

**Conclusion:** Solitary plasmacytoma commonly affects young males. Bones are more commonly affected than extramedullary sites. SP has a lower rate of progression and excellent prognosis when treated with local radiotherapy.

## Introduction

Solitary plasmacytomas (SP) are rare plasma cell neoplasms which comprise 5-6% of plasma cell disorders (1,2). It is a localized involvement by clonal plasma cells and is a distinct clinical entity from multiple myeloma (MM). Depending on whether bone or extramedullary site is involved, it can be classified into solitary bone plasmacytoma (SBP) or solitary extramedullary plasmacytoma (SEP). The current classification of SP also includes an entity with minimal bone marrow involvement also known as Plasmacytoma +, which is characterized by <10% clonal plasma cells in bone marrow (3). SBP mostly affects bones of the axial skeleton like vertebrae and skull (4,5). SEP mostly affects the head and neck region, most frequently nose and nasopharynx (6,7). It commonly affects males and the median age at presentation is 55 to 60 years (4,5,7).

Traditionally, the mainstay of treatment of solitary plasmacytomas with and without bone marrow involvement has been localized radiotherapy in most cases or surgical excision in selected instances especially in SEP. Initial treatment is followed by regular follow up as some patients will progress to SP or MM (8–10). There are varied prognostic indicators of disease progression to SP or MM such as bone or extramedullary involvement, minimal bone marrow (BM) involvement, persistent M protein or abnormal free light chain (FLC) after localized therapy (11). Apart from these traditional risk factors, presence of high risk cytogenetic abnormality (HRCA) and mutations involving MAPK pathway, KRAS, TP53 and genes involving DNA damage repair (ATM, ATR, DIS3) is associated with shorter progression free survival (12,13).

Since there is very limited data on solitary plasmacytoma from India, the purpose of this study was to describe the clinical characteristics, treatment patterns, outcomes and prognostic factors of disease progression.

## Materials and Methods

### Study Population

We retrospectively reviewed the medical records of solitary plasmacytoma (SBP and SEP) from electronic medical records (EMR) from January 2012 to December 2025. Patients who were diagnosed before 2012 in an outside center, were also included provided the patients were on regular follow up. Patients with more than one plasmacytoma, SP with >10% clonal plasma cells in bone marrow, SP fulfilling the criteria of smouldering myeloma (SMM) or multiple myeloma (MM), POEMS (polyneuropathy, organomegaly, endocrinopathy, monoclonal protein, skin changes) syndrome has been excluded from analysis.

Relevant clinical, demographic and laboratory data were extracted from EMR. The variables included, age, gender, chief complaints, performance status, hemoglobin, calcium, creatinine, M band, serum immunofixation electrophoresis, serum free light chain ratio, immunoglobulin levels, beta2 microglobulin, bone marrow aspiration, trephine biopsy, plasma cells clonality assessment by immunohistochemistry (IHC) of flow cytometry, cytogenetic abnormality by fluorescent in situ hybridization (FISH), histopathological examination and IHC on biopsy block, MRI or PET CT finding, treatment received were included. Bone marrow aspirate, trephine biopsy and tissue block done elsewhere were reviewed prior to therapy initiation. Prior to data collection, approval from the institutional review board (IRB) was taken. Patient confidentiality and privacy were protected through de-identification of the data from the EMR. The study conforms to the declarations of Helsinki (14).

### Statistical analysis

Data were analyzed using SPSS v31 and R statistical software. Descriptive statistics and categorical variables were presented as frequencies and percentages. Survival outcomes including progression free survival (PFS), overall survival (OS), time to next treatment (TTNT) estimated by kaplan meier method. PFS is calculated from the time of diagnosis to progression to myeloma, solitary plasmacytoma, biochemical progression leading to treatment initiation and death. OS was calculated from the date of diagnosis to the last follow up date or date of death. TTNT was calculated as the time interval between first line and second line treatment. Differences in survival curves between subgroups were assessed with a log-rank test. Cox regression analysis was used for univariate and multivariate analysis. A p-value < 0.05 was considered statistically significant.

## Results

### Baseline characteristics

46 patients of solitary plasmacytoma registered between January 2012 and December 2025. The median age of the studied population was 53 years (23-75 years). Males were more commonly affected than females (M:F=2.28:1). Most common chief complaints were bony pain (67.4%) followed by localised swelling (17.4%), paraplegia (6.5%), cough (2.2%), dyspepsia (2.2%), nasal blockade (2.2%) and quadriplegia (2.2%). SBP was seen in 39 (84.8%) cases whereas SEP was seen in 7 (15.2%) cases ***(Table 1)***. The most common site of SBP was vertebra (61.5%) followed by skull (7.7%), sternum (5.1%), clavicle (5.1%), pelvic bones (5.1%), sacrum (5.1%), orbit (2.56%), scapula (2.56%), femur (2.56%), and fibula (2.56%) whereas the most common site of SEP was lung (28.57%) followed by pituitary gland (14.28%), nose (14.28%), Thyroid gland (14.28%), Lymph node (14.28%) and Stomach (14.28%).

**Table 1:** Baseline characteristics.

|  |  |  |
| --- | --- | --- |
| Characteristics |  | n=46 (%), |
| Age in years, Median (range) |  | 53 (23-75) |
| Gender | Male | 32 (69.6%) |
|  | Female | 14 (30.4%) |
| Hb (g/dl) median, range |  | 12.7 (8.6-16.5) |
| Calcium (mg/dl), median, range |  | 8.9 (6.9-10.1) |
| Creatinine (mg/dl), median, range |  | 0.71 (0.32-1.5) |
| Presence of M band |  | 25 (54.3%) |
| M band (g/dl), Median (range) |  | 0.24 (0.10-1.95) |
| Isotype (n=33) | IgG | 20 (60.6%) |
|  | IgA | 5 (15.1%) |
|  | Light Chain | 4 (12.1%) |
|  | Negative | 4 (12.1%) |
| Involved:Uninvolved Light chain ratio (mg/L), Mean, range |  | 3.03 mg/L (0.36-36.31) |
| Site | Solitary Bone (SBP) | 39 (84.8%) |
|  | Solitary Extramedullary (SEP) | 7 (15.2%) |
| Bone marrow involvement |  | 6 (13%) |
| Cytogenetics (n=6) | 17p del | 1 (16.6%) |
|  | t (11;14) | 1 (16.6%) |
|  | Negative | 4 (66.67%) |
| Light Chain restriction on biopsy (n=44) | Kappa | 26 (59%) |
|  | Lambda | 18 (41%) |

**Table 2:** Anatomic location of solitary bone plasmacytoma and solitary extramedullary plasmacytoma.

| Solitary Bone plasmacytoma | n=39 (%) | Solitary Extramedullary Plasmacytoma | n=7 (%) |
| --- | --- | --- | --- |
| Vertebra | 24 (61.5%) | Lung | 2 (28.57%) |
| Skull | 3 (7.7%) | Pituitary | 1 (14.28%) |
| Sternum | 2 (5.1%) | Nose | 1 (14.28%) |
| Clavicle | 2 (5.1%) | Thyroid | 1 (14.28%) |
| Pelvic bones | 2 (5.1%) | Lymph Node | 1 (14.28%) |
| Sacrum | 2 (5.1%) | Stomach | 1 (14.28%) |
| Orbit | 1 (2.56%) |  |  |
| Scapula | 1 (2.56%) |  |  |
| Femur | 1 (2.56%) |  |  |
| Fibula | 1 (2.56%) |  |  |

### Laboratory characteristics

All the cases were confirmed with a histopathological examination on a local biopsy or excised surgical sample. Immunohistochemistry was used to confirm the diagnosis using anti CD138 and CD56 antibodies. Clonality was assessed with anti kappa and lambda stains in 44 cases. Out of 44 cases, kappa restriction was seen in 26 (59%) cases whereas lambda restriction was seen in 18 (41%) cases. Median Hemoglobin, calcium and creatinine was 12.7 g/dL (8.6 to 16.5 g/dL), 8.9 mg/dL (6.9-10.1 mg/dL) and 0.71 mg/dL (0.32-1.5 mg/dL) respectively. M band was present in 25 cases (54.3%). Median M band concentration 0.24 g/dL (0.1 to 1.95 gm/dL).

Immunofixation electrophoresis was done in 33 cases. IgG was the most common isotype accounting for 60.6% cases followed by IgA (15.1%) and Light chain (12.1%). Mean involved:uninvolved light chain ratio was 3.03 mg/L (0.36-36.31 mg/L). It was negative in 4 cases. Bone marrow studies were done in all cases and only 6 (13%) cases showed involvement by clonal plasma cells. Cytogenetic abnormality was assessed by Fluorescence in situ hybridization (FISH) on CD138 sorted plasma cells from bone marrow samples where clonal plasma cells were detected. One patient had 17p deletion, one patient had t(11;14) and remaining cases were negative. In remaining patients FISH analysis was not done as there was no involvement by clonal plasma cells. ***(Table 1)***

### Treatment

In the whole cohort, 41 (89.1%) patients received radiotherapy alone or in combination with chemotherapy. Median dose received was 45 Gy (20-55 Gy) 36 (78.2%) patients received only radiotherapy whereas 5 (10.9%) patients received radiotherapy in combination with systemic therapy. In the systemic therapy cohort two (4.3%) patients received Bortezomib Lenalidomide Dexamethasone (VRD), one (2.17%) patient each received Bortezomib Adriamycin Dexamethasone (VAD), Cyclophosphamide Thalidomide Dexamethasone (CTD) and Thalidomide Dexamethasone (TD) respectively. Two patients received maintenance for two years with Lenalidomide. Five (10.9%) patients underwent total surgical excision with no further radiotherapy or systemic therapy. Post treatment these patients were on close observation with biochemical tests. In the entire cohort, 14 (30.4%) patients experienced disease progression. Three (6.5%) patients experienced recurrent SBP in a new site, 11 (24%) patients progressed to MM. Two (4.34%) patients died in the entire cohort. Both the deaths are unrelated to plasma cell neoplasm. ***(Table 3)***

**Table 3:** Treatment and outcome.

|  |  |  |
| --- | --- | --- |
| Treatment |  | n=46(%) |
| Radiotherapy |  | 36 (78.2%) |
| Surgery |  | 5 (10.9%) |
| Radiotherapy and Systemic Therapy (n=5) | Bortezomib Lenalidomide Dexamethasone (VRD) | 2 (4.3%) |
|  | Bortezomib Adriamycin Dexamethasone (VAD) | 1 (2.17%) |
|  | Cyclophosphamide Thalidomide Dexamethasone (CTD) | 1 (2.17%) |
|  | Thalidomide Dexamethasone (TD) | 1 (2.17%) |
| Disease progression (n=14) | Solitary Plasmacytoma | 3 (6.5%) |
|  | Multiple Myeloma | 11 (23.9%) |
|  | Total | 14 (30.4%) |
| Death |  | 2 (4.34%) |

### Survival analysis

With a median follow up of 5.4 years (95% CI: 1.8 – 9.0), median OS was not reached, median PFS was 9.22 years (95% CI: 5.8-12.6), median time to next treatment (TTNT) was 9.86 years (95% CI: 6.8 – 12.9) (***Figure 1)***. Median PFS of SBP and SEP was 8.8 years (95% CI: 5.7-12.0) and 9.2 years (6.6-11.7). Factors that influence progression in the current cohort were analysed with a univariate cox regression analysis. With respect to site (SBP vs SEP), bone marrow involvement, presence of M band, abnormal involved to uninvolved FLC ratio, use of systemic anti-myeloma therapy have not demonstrated any prognostic impact. (***Table 4)***

**Table 4:** Univariate analysis of baseline factors and progression free survival.

| Variable | Univariate Analysis |  |  |
| --- | --- | --- | --- |
|  | P value | Hazard Ratio (HR) | 95% Confidence Interval (CI) |
| SBP vs SEP | 0.63 | 1.64 | 0.21-12.7 |
| Bone marrow Involvement | 0.45 | 0.61 | 0.17-2.2 |
| Presence of M band | 0.62 | 1.30 | 0.44-3.7 |
| Abnormal Involved/Uninvolved Ratio | 0.89 | 0.92 | 0.26-3.2 |
| Systemic anti- | 0.29 | 3.0 | 0.37-23.9 |
| myeloma Therapy |  |  |  |

**Fig 1:**
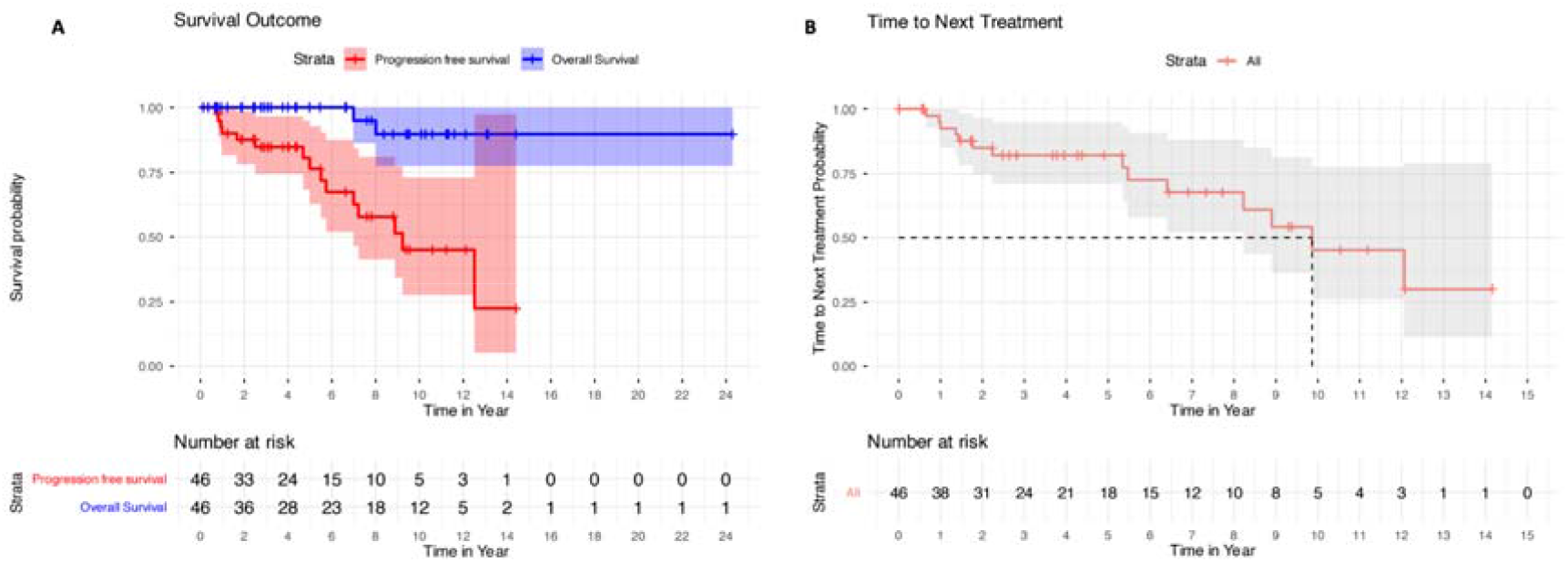
Overall survival and Progression Free survival, 1B: Time to next treatment

## Discussion

Solitary plasmacytomas are rare diseases characterised by localised proliferation of clonal plasma cells. With the advent of more sensitive techniques like Immunohistochemistry, Flowcytometry on Bone marrow samples, MRI and PET CT scan, systemic involvement can be easily ruled out. SP has a favourable prognosis when compared to MM.

SP affects relatively older patients with a median age of 60 years. The current study reports a slightly lower median age of 53 years (8,15). Previous studies from India also reported a lower median age of 49 to 56.7 years (16,17). Males are more commonly affected than females with a M:F ratio of 2.28:1, which is similar to previous reported studies by Basavaiah et al (M:F=1.9:1) and Baghmar et al (M:F=1.8:1) (16,17). SBP (84.8%) are more common than SEP (15.2%). These findings are also similar to previous studies where bone involvement was 86.6% (Baghmar et al), 90.5% (Charalampous et al) whereas slightly lower bone involvement was seen in studies by Basavaiah et al (57%), Katodritou et al (67%) and Dores et al (58%) (2,8,15–17).

Among patients with SBP, most common anatomic site involvement was the vertebral column (61.5%) followed by skull (7.7%). Vertebral column involvement is also seen in other studies by Katodritou et al (40%), Basavaiah et al (31.2%), and Baghmar et al (42%) (15–17). Among the extramedullary sites, the current study reports a higher incidence of lung involvement (28.57%) followed by one case each of nasal, thyroid, stomach, lymph node and pituitary gland. Most common site for SEP in other studies are nasal cavity (37.5%), paranasal sinus (50%), nose and nasopharynx (53.12%) as reported by Baghmar et al, Basavaiah et al and Katodritou et al respectively (15–17).

Presence of serum M band was seen in 54.3% in the current study is also similar to other studies by Charalampous et al (60%), Baghmar et al (48%) and Basavaiah et al (41.7%) whereas higher M band positivity was seen in studies by Katodritou et al (80%) (8,15–17). Immunofixation electrophoresis was used to determine the Immunoglobulin (Ig) Isotype in 33 cases. The most common Ig Isotype in the current study is IgG (60.6%) followed by IgA (15.1%), Light Chain (12.1%) and Negative (12.1%). The most common Isotype reported by Dingli et al is as follows: IgG in 49%, IgA in 9%, Light chain in 3%, IgM in 1% Biclonal in 2% and Negative in 36% cases (11).

Clonal Plasma cells in bone marrow (Solitary Plasmacytoma with Minimal marrow Involvement, Plasmacytoma +) were seen in 13% cases. Incidence of plasmacytoma with minimal marrow involvement varies from 38% to 48% in the published literature (8,18). The higher incidence of Plasmacytoma + in other cohorts can be explained by the method of assessment of clonal plasma cells. In the current study, we used Immunohistochemistry on trephine biopsy whereas in studies by Katodritou et al and Charalampous et al used Flowcytometry as well as immunohistochemistry on trephine biopsy for assessment of clonal plasma cells. Since only six patients had bone marrow involvement, FISH was done in 6 patients. One patient each had 17p deletion and t (11;14). The remaining four cases were negative. Cytogenetic abnormalities have been reported to be as high as 48% with high risk cytogenetic abnormalities comprising almost 19-20% of the entire cohort (13). The lower incidence of cytogenetic abnormality in the current study is probably because of lack of use of Interphase FISH on tissue blocks and not incorporating flowcytometry at diagnosis.

Local radiotherapy is the cornerstone of management of SBP and complete surgical excision is the mainstay of management of SP which are not amenable to radiotherapy. In the current cohort, 78.2% patients received radiotherapy, 10.9% each patient received surgical excision and radiotherapy with anti-myeloma therapy respectively. Median radiotherapy dose received was 45 Gy (20-55Gy). This finding is similar to previous reported studies where radiotherapy was the main modality of management (8,16,18). Although the use of concurrent systemic anti-myeloma therapy did not lead to improved PFS in the current study, there is data to show that use of systemic therapy decreases the likelihood of progression to MM (19). Currently, the IDRIS trial (IDRIS Trial, clinicaltrials gov. Identifier: NCT02544308) is evaluating the role of Lenalidomide in high risk solitary plasmacytoma.

In the current study, 14 patients progressed to SP or MM in the entire follow up duration. With a median follow up of 5.4 years, median OS was not reached, median PFS was 9.22 years. This is in line with previously reported studies, where median OS ranged from 15.2 to 18.7 years and median progression free survival ranged from 2.5 years to 10 years. Charalampos et al did a meta analysis and review of 62 studies and they found that two-third patients were disease free at 3 years and and more than half (55%) patients were disease free at 5 years (8).

With respect to baseline risk factors affecting progression free survival, multiple studies have shown that patients with SBP, abnormal FLC, presence of clonal plasma cells in bone marrow, use of systemic antimyeloma therapy has shorter progression free survival (8,13,18). However Baghmar et al did not find any correlation between these factors and high PFS (16).

High risk cytogenetic abnormality has been associated with shorter PFS (13). Incorporating FISH on the diagnostic biopsy block is necessary to ascertain cytogenetic abnormality. The current study used FISH analysis only on bone marrow aspirates which showed increased proportion of plasma cells and therefore could only identify one patient with deletion 17p.

The limitations of this study are its retrospective nature, small sample size, limited use of flow cytometry for assessment of bone marrow involvement, limited assessment of cytogenetic abnormalities by FISH on the diagnostic biopsy sample and absence of immunoelectrophoresis in all cases.

To conclude, solitary plasmacytoma commonly affects relatively young males. Bones are more commonly affected than extramedullary sites. SP has a lower rate of progression and excellent prognosis when treated with local radiotherapy.

## Data Availability

All data produced in the present study are available upon reasonable request to the authors

